# Using the infection fatality rate to predict the evolution of Covid-19 in Brazil

**DOI:** 10.1101/2020.07.01.20144279

**Authors:** M. S. Cecconello, G. L. Diniz, E. B. Silva

**Affiliations:** Department of Mathematics - ICET - UFMT; Department of Statistics - ICET - UFMT

**Keywords:** IFR, SIR model, epidemiological model

## Abstract

In this work we are going to use estimates of Infection Fatality Rate (IFR) for Covid-19 in order to predict the evolution of Covid-19 in Brazil. To this aim, we are going to fit the parameters of the SIR model using the official deceased data available by governmental agencies. Furthermore, we are going to analyse the impact of social distancing policies on the transmission parameters.

## 1 Introduction

Currently, the world is experiencing an unprecedented serious pandemic crisis, which has led to the collapse of health systems in several countries, with the aggravating lack of hospital beds and individual protection material, as well as insufficient testing supplies for the diagnosis of patients. In this sense, the data of infected patients does not always correspond to reality, since there is a much larger number of cases not reported by governmental health agencies, which is called under-reporting. The estimate of under-reporting in most countries has been 10 for each patient notified, based on the fatal infected rate (IFR) compared to this rate before the pandemic was established (Ferguson et al., 2020). Due to the limited information available, most parameters describing the dynamics of the disease spread involve significant uncertainties (Lachmann et al., 2020). Healthcare systems in most countries are not capable of monitoring the exponential growth of a virus in this manner. South Korea, as of writing, has one of the most extensive capabilities of testing individuals per capita, with a capacity of more than 20,000 tests a day. Hence, South Korea represents the best benchmark country in order to predict the COVID-19 CFR Lachmann et al. (2020). Another important factor to be considered is the effect of social distancing on the value of the transmission parameter of the disease, which is not considered in classic epidemiology models. This is the relevant aspect of the model that we are considering in this paper.

Severe acute respiratory syndrome coronavirus (SARS-CoV) and Middle East respiratory syndrome coronavirus (MERS-CoV) are two highly transmissible and pathogenic viruses that emerged in humans at the beginning of the 21st century (Cui et al., 2018). The recently emerged Severe Acute Respiratory Syndrome Coronavirus 2 (SARS-CoV-2) pandemic, with first cases reported in Wuhan, China in late December, 2019, quickly spread to other countries and was declared by the World Health Organization (WHO) as of January 30, 2020, an Public Health Emergency of International Concern (PHEIC). PHEIC are extraordinary events which pose a large scale public health risk with international spreading and which, in general, require a coordinated response. In Brazil, national public health emergencies (NPHE) are defined according to Brazilian Ministry of Health (MoH) as events that represent risks to public health and that occur in situations of outbreaks or epidemics (as a result of unexpected agents or reintroduction of eradicated diseases or with high severity), disasters and of lack of assistance to the population, which go beyond the response capacity of the state (Croda et al., 2020).

The coronavirus 2019 (COVID-19) pandemic has been spreading globally for months, yet the infection fatality ratio (IFR) of the disease is still uncertain. This is partly because of inconsistencies in testing and death reporting standards across countries (Rinaldi and Paradisi, 2020). IFR is the ratio of deaths divided by the number of actual infections with SARS-CoV-2 (Condit, 2020). However for the coronavirus 2019 (COVID-19) infection with its broad clinical spectrum from asymptomatic to severe disease courses, the IFR is the more reliable parameter to predict the consequences of the pandemic. Case fatality rate (CFR) is the proportion of the number of deaths divided by the number of confirmed patients of a disease, which has been used to assess and compare the severity of the epidemic between countries. The rates can also be used to assess the healthcare capacity in response to the outbreak (Kim et al., 2020). According to Condit (2020), the IFR is likely to be significantly lower than the CFR because usually only people with severe symptoms are being tested but exist a large number of infections with SARS-CoV-2 result in mild or even asymptomatic disease.

The first case reported in Brazil was on February 26, 2020, in the city of São Paulo, of a 61-year-old man who had made a trip to Italy at that time SanarMED (2020). Until May 20, the state of São Paulo recorded 5,363 deaths from the new coronavirus, with 216 deaths confirmed in the last 24 hours. The state also totals 69,859 confirmed cases of COVID-19, with one or more people infected in 484 cities. At least one fatal victim was recorded in 223 municipalities. Another contribution to the understanding of this pandemic has been made by our research group Diniz (2020), in order to analyze the number of confirmed cases and number of deaths, proportionally the population of each state, that is necessary to view these number in the same scale, avoiding the serious mistake of comparing the absolute numbers. This methodology allowed to make a ranking with all the federate units, including Brazil that served for the national average.

In order to mitigate the effects of the pandemic in the health system, governments have been adopting several policies of social distancing world-wide. In general, these social distancing policies range from just avoiding overcrowding, to closing school, bars, restaurants and so on to complete lock-down. Furthermore, some countries and cities have been adjusting their social distancing policies over time, depending on to the pressure over the health system. On the other hand, people not always follows the rules imposed by governments. With this in mind, we are going to assume that the fluctuations on the levels of social distancing policies can be analyzed looking at changes in the transmission parameter of the SIR model. In order to analyze this changes, we are going to split the data into disjoint subsets, fit the model to the data in each one of this subset and carry out an analysis of variance on the parameters.

## 2 The SIR model

A classic mathematical model to describe the spread of an infectious disease on a population as function of time is the SIR model. In this model, individuals of a population are labeled in three categories: susceptible (S), infected (I) and recovered (R). Considering the proportion of each category in relation to the total population, such proportions evolve in time according to the following equation

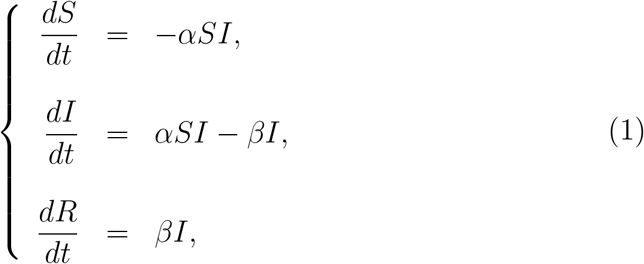

in which *α* and *β* are both positive parameters Edelstein-Keshet (2005).

In the equation (1), parameter *α* is responsible for how often a susceptible-infected interactions results in a new infection by units of time. Thus, *α* proportional to the probability of transmitting disease between a susceptible and an infectious individual. Yet *β*, is a recovery rate, measured by units of time, so that 1*/β* is the the average duration of infection in the organism of individuals.

The behavior of SIR model solution depends on the values of *α* and *β*, as well as in the initial proportions *S*_0_, *I*_0_ and *R*_0_. And so, an important issue is this kind of model is in what conditions the infection spreads on the population. In order for this to happen, the derivative of *I*(*t*) must be positive, that is,

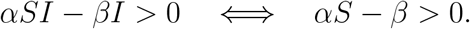

Since at the beginning almost all individuals are susceptible, then *S ≈* 1 and thus, last equation becomes

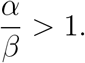

The ratio *α/β* is called *ℛ*_0_ and can be interpreted as the number of secondary infections from a single case in a completely susceptible population.

## 3 The fitting procedure and simulation study

In order to obtain the parameters to the SIR model we are going to use official data updated daily by the governmental agencies. Due to reliability, we are especially interested in the cumulative number of deaths over time. From now on, time *t* = 0 is at March 3rd of 2020 and *t* = *N* is at June 20 of 2020.

For the fitting procedure we are assuming that the recovery time *τ* is a random variable uniformly distributed on the interval [*τ*_1_, *τ*_2_] while the IFR for Covid-19 is a random variable uniformly distributed on the interval [*µ*_1_, *µ*_2_]. Therefore, if *µ ∈*[*µ*_1_, *µ*_2_] is the IFR for Covid-19 and *d*_*i*_ is the cumulative number of deaths at time *i* then, by definition, the number of recovered is given by 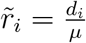.

Let *R*_*i*_ be the number of recovered at time *i* given by the solution of SIR model. As the actual number of infected are not known, to estimate the transmission parameter *α* we minimize the square error function given by

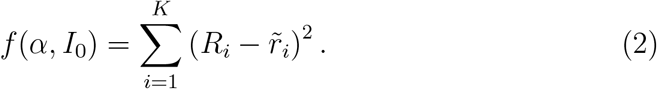

In all cases, we use *R*_0_ = *r*_0_ and *S*_0_ = 1 *− R*_0_ *− I*_0_.

As mentioned before, we are considering uncertainties in both the IFR and the time to recovery *τ* so that we randomize over these parameters. So, the fitting procedure follows the steps:

1. Considering *µ* and *τ* distributed over their domains, randomly select *µ ∈* [*µ*_1_, *µ*_2_], *τ ∈* [*τ*_1_, *τ*_2_], defining *β* = 1*/τ* and *r*_*i*_ = *d*_*i*_*/µ*;
2. Randomly select *i*_1_, *i*_2_, *…, i*_*K*_ *∈ {*0, 3, *…, N}*, allowing repetition;
3. Minimize function 2 using the data 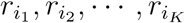.
4. The steps 1 *−* 3 are repeated *M* times.

For each one of the *M* iterations we set *S*_0_ = 1 *− I*_0_ *− R*_0_ and *R*_0_ = *r*_0_. In the minimizing step 3, we are using the *fminsearch* function from Matlab, and thus, we consider the minimizing steps reaches a local minimum in cases in which the options *Tolfun* and *TolX* are less than 1*e −* 5.

### 3.1 Capturing social distancing effects

As described in the Introduction, we want also to capture the effects of social distancing over time. In this case, we are assuming that social distancing policies confer changes in the value of the transmission parameter *α* and, consequently, in the reproductive number of the disease. Thus, looking at different subsets of the data are going to produce a different fitting and we can compare the changes in the parameters.

In this case, we adapt the previous fitting procedure performing simulations as the following:

1. Considering *µ* and *τ* distributed over their domains, randomly select *µ ∈* [*µ*_1_, *µ*_2_], *τ ∈* [*τ*_1_, *τ*_2_], defining 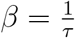 and 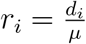;
2. For each *i ∈ {*1, 2, *…, N −* 1*}*, minimize the function 2 using the data *r*_*i−*1_, *r*_*i*_ and *r*_*i*+1_, setting iterations we set *S*_0_ = 1 *− I*_*i*_ *− R*_*i*_ and *R*_0_ = *r*_*i*_.
3. The steps 1 *−* 2 are repeated *M* times.

Each one of these *M* simulations produces a value of *α* at some *t* = *i, i ∈ {*1, 2, *…, N −* 1*}*. That is, each of these *M* simulations finds a SIR solution that approaches better to the data *r*_*i−*1_, *r*_*i*_ and *r*_*i*+1_. Thus, in order to compare these values we calculate the reproductive number *ℛ*_*t*_ at *t* = *i* by

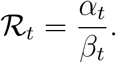

Next section we are going to use the procedures describe in this section in order to analyze the dynamics of the pandemic.

### 3.2 Analysis of the simulations

To analyse the results obtained in the simulations we calculate the averages of the estimates and the confidence intervals (95%), the root-mean-square errors and the empirical standard error (ESE). For ***θ*** = (*α, ℛ*_*t*_), the simulated root-mean-square errors for the *M* simulations is obtained from the expression

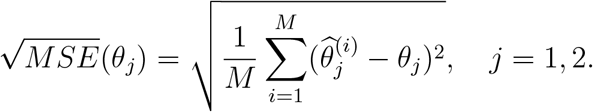

The empirical standard errors (ESE) are obtained from the root of variance for the *M* simulations given by

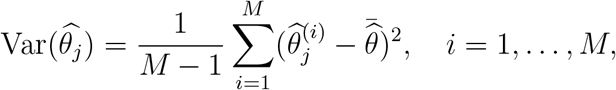

in that 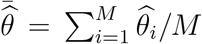 and 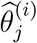 represents the estimated of *θ*_*j*_ in the *i*-th simulation.

## 4 Results

In this section, we present and discuss the results obtained using the methods described in last section.

### 4.1 Under-reporting

To the results of this section we set *M* = 100 and *K* = 65, which corresponds to 75% of the data available. Furthermore, we consider 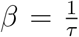 in which *τ* is a random variable uniformly distributed over [9, 19] and *µ* is a random variable uniformly distributed over [0.005, 0.01].

To measure the under-reporting we consider the mean simulation results for cumulative cases as described in Section 3 and calculate the ratio over the official cumulative cases reported by governmental agencies. That is, we have been considering *C*_*i*_ = *R*_*i*_ + *I*_*i*_ the mean cumulative cases predicted by the SIR model and *c*_*i*_ the reported number of total cases, we compute the under-reporting ratio by

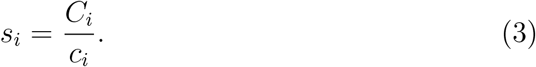

In Figure 1 we can see the evolution of the under-reporting ratio over time.

**Figure 1:**
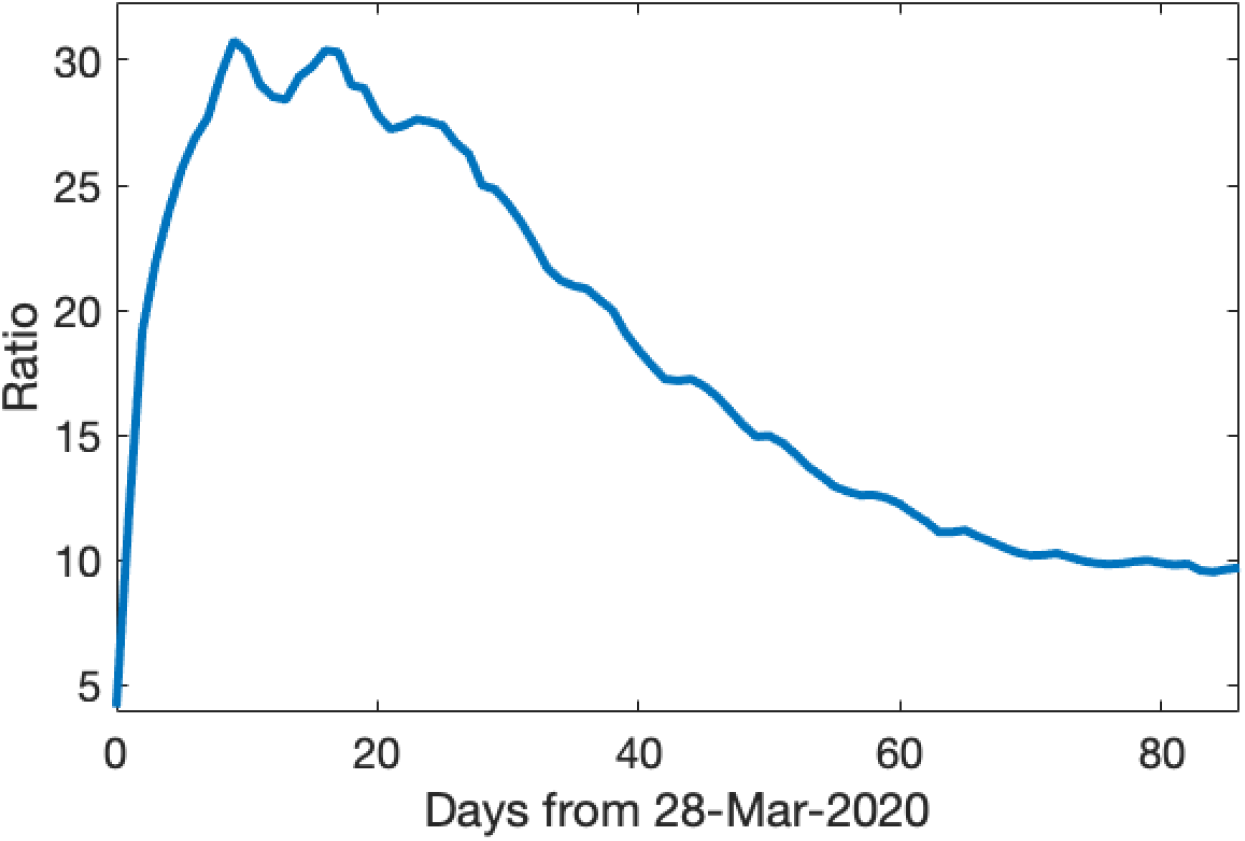
Under-reporting over time.

Using the expression given in equation (3), we calculate the daily underreporting rates for Brazil considering the period from March 28 to June 22. With these values, we generate Figure 1 and calculate the average underreporting for Brazil that was approximately 18.16 with a standard deviation of approximately 7.52 and a coefficient of variation of approximately 41.39%. That is, each reported case corresponds to a 1*/*18.16 of the actual number of people that have contact with the virus.

However, it is worthy to note that the under-reporting have been decreasing in last days. In Figure 3 we can see the under-reporting ratio is about 10 (9.78 is the average in last 10 days) meaning that each reported case represent (1/10) of actual of people that have contact with the virus.

### 4.2 Social distancing in Brazil

In this section we present the results for reproductive number *ℛ*_*t*_ and six distance measurements available at Google LLC (2020).

To the results of the reproductive number *ℛ*_*t*_ of this section we set

*M* = 10000 and *K* = 65, which corresponds to 75% of the data available.

Furthermore, we consider 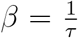 in which *τ* is a random variable uniformly distributed over [9, 19] and *µ* is a random variable uniformly distributed over [0.005, 0.01].

The data available at Google LLC (2020) contain movement trends over time by geography, across different categories of places such as retail and recreation, groceries and pharmacies, parks, transit stations, workplaces, and residential. These measurements are:

*M*_1_ : Mobility trends for places like restaurants, cafes, shopping centers, theme parks, museums, libraries, and movie theaters.

*M*_2_ : Mobility trends for places like grocery markets, food warehouses, farmers markets, specialty food shops, drug stores, and pharmacies.

*M*_3_ : Mobility trends for places like national parks, public beaches, marinas, dog parks, plazas, and public gardens.

*M*_4_ : Mobility trends for places like public transport hubs such as subway, bus, and train stations.

*M*_5_ : Mobility trends for places of work.

*M*_6_ : Mobility trends for places of residence.

The measurements (*M*_*i*_, *i* = 1, …, 6) shows how visits and length of stay in different locations change compared to a reference value. The changes for each day are compared with a reference value corresponding to the same day of the week. The reference value is the median of the corresponding day of the week, during the period of five weeks from January 3 to February 6, 2020 (Google LLC, 2020).

According to Google LLC (2020), these measurements is intended to be used to mitigate the impact of Covid-19. It shouldn’t be used for medical diagnostic, prognostic, or treatment purposes. It also isn’t intended to be used for guidance on personal travel plans. Each Community Mobility Report data-set is presented by location and highlights the percent change in visits to places like grocery stores and parks within a geographic area. How to use this report. Location accuracy and the understanding of categorized places varies from region to region, so it isn’t recommended using this data to compare changes between countries, or between regions with different characteristics (e.g. rural versus urban areas).

To seek for a correlation between those measurements and their effects on the dynamics of the epidemic in Brazil we group the data by week considering average values. Thus, the first column in 1 shows the week number using 2020 as the reference year. Yet the values of *ℛ*_*t*_ are average values of the *ℛ*_*t*_ values, obtained by using the procedure described in subsection 3.1, in which the *ℛ*_*t*_ values are grouped by week. similarly, each *M*_*i*_ value shown in Table 1 represents a weekly averages of the values generated by the site Google LLC (2020).

**Table 1:**
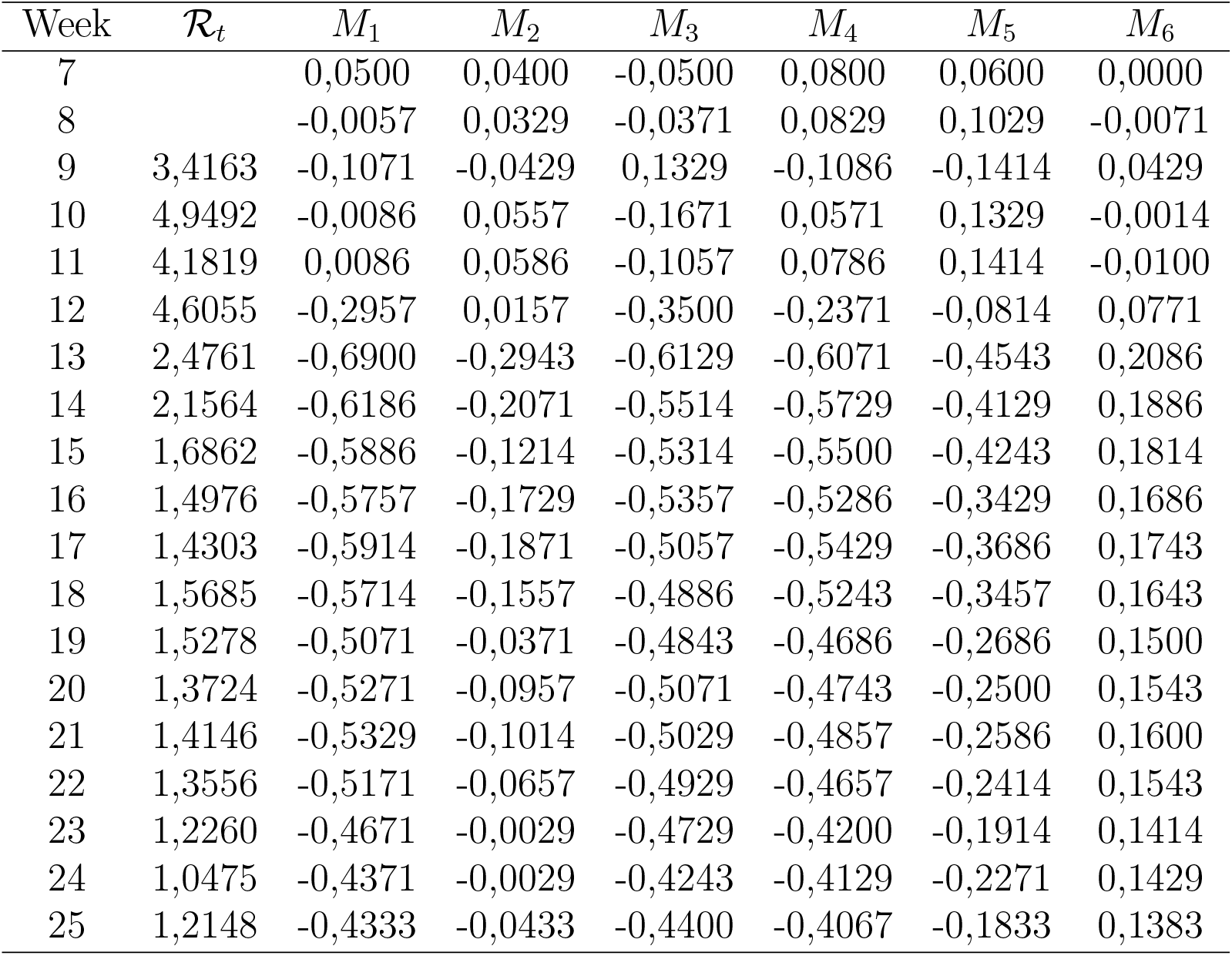
Weekly average values for the reproductive number *ℛ*_*t*_ and Google social distancing measurements.

| Week | $\mathcal{R}_t$ | $M_1$ | $M_2$ | $M_3$ | $M_4$ | $M_5$ | $M_6$ |
| --- | --- | --- | --- | --- | --- | --- | --- |
| 7 |  | 0,0500 | 0,0400 | -0,0500 | 0,0800 | 0,0600 | 0,0000 |
| 8 |  | -0,0057 | 0,0329 | -0,0371 | 0,0829 | 0,1029 | -0,0071 |
| 9 | 3,4163 | -0,1071 | -0,0429 | 0,1329 | -0,1086 | -0,1414 | 0,0429 |
| 10 | 4,9492 | -0,0086 | 0,0557 | -0,1671 | 0,0571 | 0,1329 | -0,0014 |
| 11 | 4,1819 | 0,0086 | 0,0586 | -0,1057 | 0,0786 | 0,1414 | -0,0100 |
| 12 | 4,6055 | -0,2957 | 0,0157 | -0,3500 | -0,2371 | -0,0814 | 0,0771 |
| 13 | 2,4761 | -0,6900 | -0,2943 | -0,6129 | -0,6071 | -0,4543 | 0,2086 |
| 14 | 2,1564 | -0,6186 | -0,2071 | -0,5514 | -0,5729 | -0,4129 | 0,1886 |
| 15 | 1,6862 | -0,5886 | -0,1214 | -0,5314 | -0,5500 | -0,4243 | 0,1814 |
| 16 | 1,4976 | -0,5757 | -0,1729 | -0,5357 | -0,5286 | -0,3429 | 0,1686 |
| 17 | 1,4303 | -0,5914 | -0,1871 | -0,5057 | -0,5429 | -0,3686 | 0,1743 |
| 18 | 1,5685 | -0,5714 | -0,1557 | -0,4886 | -0,5243 | -0,3457 | 0,1643 |
| 19 | 1,5278 | -0,5071 | -0,0371 | -0,4843 | -0,4686 | -0,2686 | 0,1500 |
| 20 | 1,3724 | -0,5271 | -0,0957 | -0,5071 | -0,4743 | -0,2500 | 0,1543 |
| 21 | 1,4146 | -0,5329 | -0,1014 | -0,5029 | -0,4857 | -0,2586 | 0,1600 |
| 22 | 1,3556 | -0,5171 | -0,0657 | -0,4929 | -0,4657 | -0,2414 | 0,1543 |
| 23 | 1,2260 | -0,4671 | -0,0029 | -0,4729 | -0,4200 | -0,1914 | 0,1414 |
| 24 | 1,0475 | -0,4371 | -0,0029 | -0,4243 | -0,4129 | -0,2271 | 0,1429 |
| 25 | 1,2148 | -0,4333 | -0,0433 | -0,4400 | -0,4067 | -0,1833 | 0,1383 |

By the dynamics of the reporting process and processing data from by authorities, we are assuming that there is a time-lag between time of the infection. That is, fluctuations in the measurements *M*_*i*_ are going to affect the reproductive number *ℛ*_*t*_ later on in time. Thus, to account for that time-lag we introduce a variable *τ ∈* [*−*2, 0], in which the lower bound *−*2 is chosen due to the data constraints in measurements *M*_*i*_.

In this way, in order to find a correlation between the values of *ℛ*_*t*_ and *M*_*i*_ we calculated the Pearson’s correlation coefficient 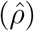 between (*ℛ*_9_, *ℛ*_10_, …, *ℛ*_25_) and 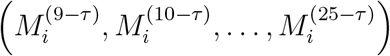 For a fixed *τ ∈* [*−*2, 0] the value of 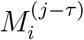. is obtained by linear interpolation. The first row in Table 2 shows the time-lag in which the maximum value for 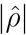 is attainable. In the second row of Table 2 we see the value of 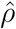 at *τ*.

**Table 2:** Pearson correlation coefficient 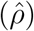 between reproductive values *ℛ*_*t*_ and the social distancing measurements from Table 1. First row shows the time delay (time-lag) between the measurement *M*_*i*_ and *ℛ*_*t*_.

| | $M_1$ | $M_2$ | $M_3$ | $M_4$ | $M_5$ | $M_6$ |
| --- | --- | --- | --- | --- | --- | --- |
| Time-lag<br>(in Weeks) | -1,4271 | -1,6583 | -1,4070 | -1,4774 | -1,5477 | -1,4673 |
| $\hat{\rho}$ | 0,8946 | 0,6039 | 0,9305 | 0,8937 | 0,8014 | -0,8917 |

According to Table 2 we see that there is a moderate to high positive linear correlation between distance measures (*M*_*i*_, *i* = 1, …, 5) and *ℛ*_*t*_, that is, as the movement in locations specified by Google LLC (2020) increases, *ℛ*_*t*_tends to increase. On the other hand, last measurement (*M*_6_) is negatively correlated with *ℛ*_*t*_. As we discuss previously, the measurement *M*_6_ refers to mobility trends for places of residence, that is, decreasing traffic on streets in residential neighborhoods there is also a decrease in *ℛ*_*t*_.

### 4.3 Epidemiological curve

One of the most important issue in epidemiological dynamics is to determine the time in which the number of infected start to decrease. In Figure 2 we show the epidemiological curve for the number of infected people. In Figure 3 we show the cumulative number of cases in proportion to the total population.

**Figure 2:**
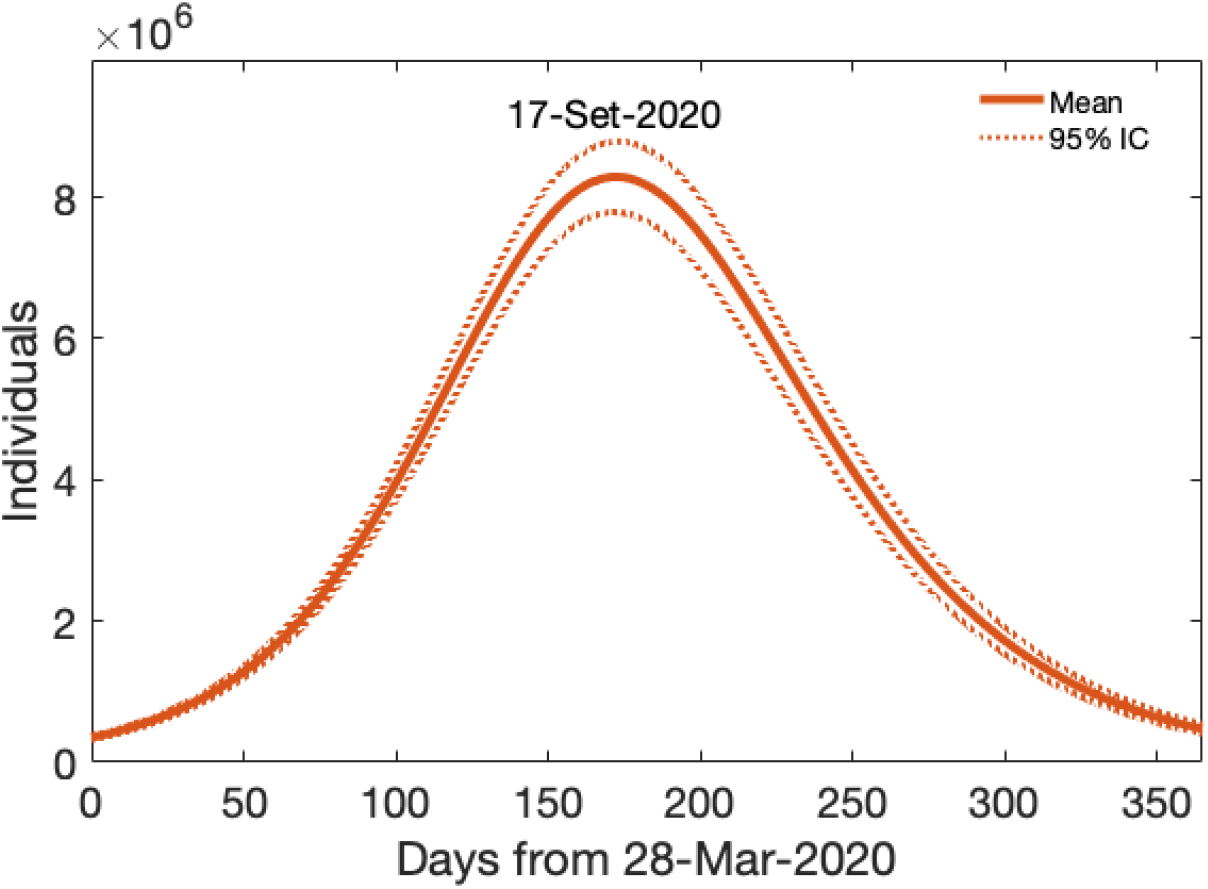
Forecast for the infectious over time. The *ℛ*_0_ is, on average, 1.36 and the standard deviation is 0.07.

**Figure 3:**
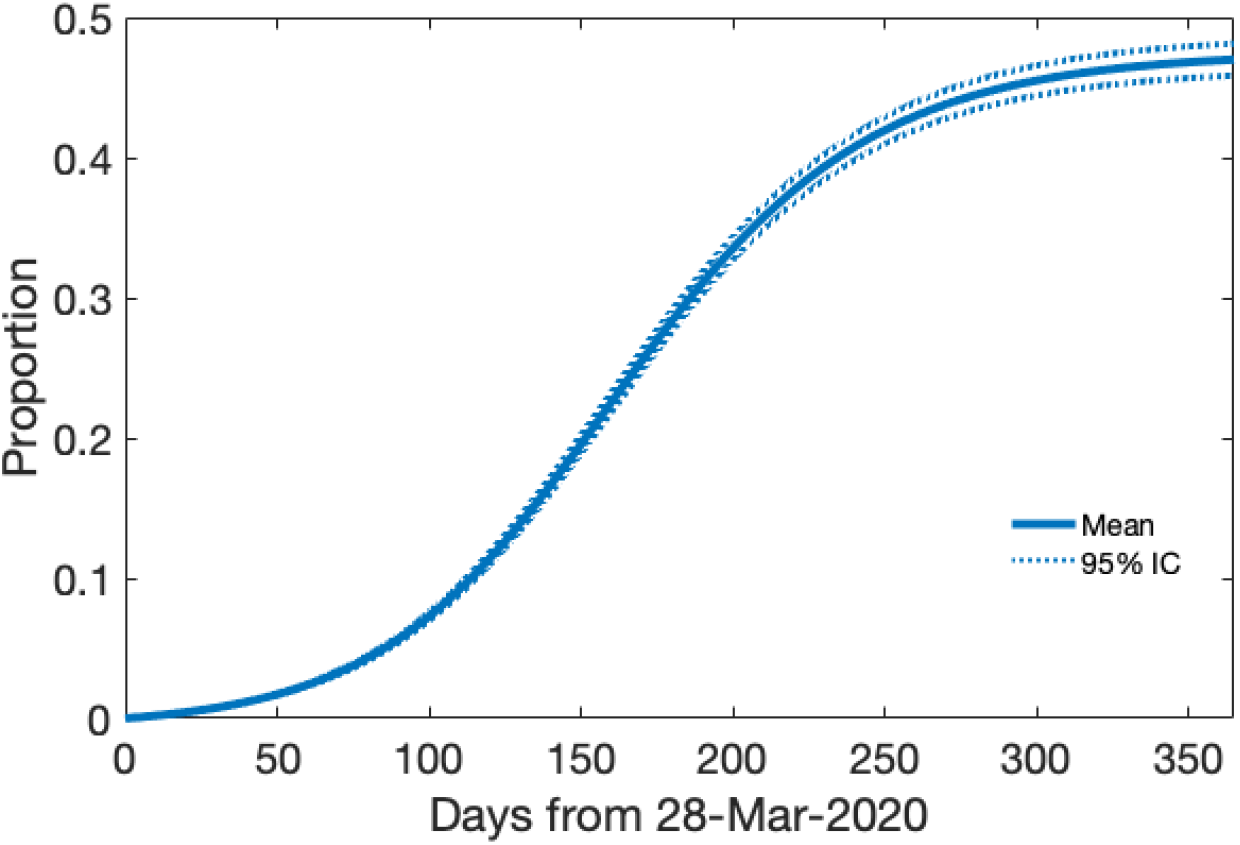
Forecast for cumulative cases over time. The *ℛ*_0_ is, on average, 1.36 and the standard deviation is 0.07.

According to the procedure adopted in this work, the number of infectious is expected to increase up to mid-September, after which the infectious start do decrease. Figure 3 shows us that about 50% of the population must be infected til April 2021.

The *ℛ*_0_ for the epidemiological curve in Figure 2 is, on average, approximately 1.36 with a standard deviation of approximately 0.07 and a coefficient of variation of approximately 5.11%

## 5 Conclusion

In this work we use the Infection Fatality Rate (IFR) in order to predict the evolution of the SARS-CoV-2 in Brazil. The main assumption to use the IFR is that we believe the accuracy of deaths is greater than the accuracy of the other data reported by governmental authorities.

By this approach we infer that the cumulative numbers of cases in Brazil is under-estimated. The ratio of under-reporting is about 18.16 meaning that each reported case corresponds to a 1*/*18.16 of the actual number of people that have contact with the virus. However, it is worthy to note that the under-reporting have been decreasing in last days. In Figure 3 we can see the that under-reporting ratio has been steady in about 10 (9.78 is the average in last 10 days) meaning that each reported case represents (1/10) of actual of number people that have contact with the virus.

Another important result in this work concerns about the effects of social distancing policies on the dissemination of the pandemic in Brazil. We infer that there is strong correlation between the reproductive number *ℛ*_*t*_ and social distancing as measured by Google LLC (2020) (presented in Table 1) in which the lowest value of *ℛ*_*t*_ occurred with the greatest adherence to social distance, and this conclusion is measured by the Pearson correlation coefficient as presented in Table 2. This is important result that can be used by authorities in order to measure how much must be the social distancing measurements in order to attain some target in the spread of the pandemic.

Finally, we believe that this approach can be helpful not just to predict the dynamics of the epidemic but also to guide policies that can control the spread of SARS-CoV-2.

## Data Availability

All data used in the manuscript are public available.

https://covid.saude.gov.br

